# Carotid-femoral pulse wave velocity is associated with post-stroke cognitive impairment

**DOI:** 10.64898/2026.05.28.26354397

**Authors:** Kevin Moncion, Lynden Rodrigues, Bernat De Las Heras, Juliano Abreu, Kira Sikorska, Adam Sutoski, Maureen J MacDonald, Ada Tang, Marc Roig

**Author notes:** **Corresponding Author:** Dr. Kevin Moncion, Memory and Motor Rehabilitation Laboratory (MEMORY-LAB), Jewish Rehabilitation Hospital, 3205 Place Alton-Goldbloom, Laval, QC H7V 1R2, Canada.

## Abstract

**Background:** Up to 70% of stroke survivors develop cognitive impairment, yet clinicians lack non-invasive vascular biomarkers that could meaningfully inform risk stratification. Carotid-femoral pulse wave velocity (cfPWV), the gold-standard measurement of central arterial stiffness, is a novel biomarker of vascular aging linked to cognitive impairment. This study evaluated the association between cfPWV and post-stroke cognitive impairment, as measured by the Montreal Cognitive Assessment (MoCA), in individuals ≥6 months post-stroke.

**Methods:** This is a secondary cross-sectional analysis of baseline data from a randomized control trial. Logistic regression analyses examined the association between cfPWV (m/s) and MoCA score at the primary cut point of ≤26/30, with secondary cut points of ≤24/30 and ≤22/30. Models were adjusted for age, sex, systolic blood pressure, type-2 diabetes, National Institutes of Health Stroke Scale (NIHSS) score, and smoking status.

**Results:** Of 82 participants enrolled in the main trial, 68 participants (n = 45 males, age 64.6 ± 9.6 years, 1.8 ± 1.2 years post-stroke) with mild-to-moderate stroke severity (NIHSS median [IQR] = 1 [2]) were included. In the fully adjusted model using the MoCA ≤26/30 cut point, each 1 m/s increase in cfPWV was associated with a 35% increase in the odds of post-stroke cognitive impairment (adjusted OR [aOR] = 1.35; 95% CI 1.06, 1.81; p = 0.027; Area Under the Curve [AUC] = 0.77). Consistent associations were observed at the MoCA ≤24/30 (aOR = 1.41; 95% CI 1.04, 2.01; p = 0.037; AUC = 0.88) and MoCA ≤22/30 (aOR = 1.33; 95% CI 1.03, 1.79; p = 0.039; AUC = 0.82) cut points.

**Conclusions:** Higher cfPWV was independently associated with post-stroke cognitive impairment across clinically referenced MoCA cut points. cfPWV may be a complementary vascular biomarker to support cognitive risk stratification and identify stroke survivors who could benefit from closer monitoring or vascular-targeted intervention.

## Introduction

Post-stroke cognitive impairment affects up to 70% of people with stroke^1-3^ and is associated with reduced independence, poorer quality of life, and increased disability and mortality.^4-8^ While cognitive deficits begin in the acute phase of stroke, many people experience a progressive, long-term deterioration in cognitive function through the chronic phases of recovery.^9,10^ There is also growing evidence that the risk of cognitive impairment post-stroke may be augmented not only by the index stroke itself,^11^ but also by the interaction of pre-existing cerebrovascular disease, traditional cardiovascular risk factors (e.g., smoking, arterial hypertension),^12^ and emerging novel risk biomarkers (e.g., blood brain barrier permeability and arterial stiffness).^11,13-15^

Arterial stiffness has emerged as a clinically relevant biomarker of vascular health^16^ that may be a precursor to poor brain health and cognitive decline in aging populations.^14,17^ Carotid-femoral pulse wave velocity (cfPWV), the gold-standard measure of central arterial stiffness,^18^ is associated with cerebral small vessel disease, microbleeds and lacunar infarcts,^19^ and white matter lesions.^20,21^ Mechanistically, stiffening of the aorta and central elastic arteries reduces their capacity to buffer pulsatile pressure and flow,^16^ exposing the high-flow and low-resistance cerebrovasculature to elevated hemodynamics.^22-24^ Indeed, these vascular changes may promote endothelial dysfunction,^25^ blood-brain barrier disruption,^26^ and adverse remodeling in the brain,^21-24,27^ all factors that contribute to accelerated cognitive decline.^14,28-30^ While associations between cfPWV and cognitive function are well-established in non-stroke populations,^28^ this relationship remains largely unexplored after stroke,^31^ particularly among a sample of individuals who are at high risk of post-stroke cognitive impairments. Thus, establishing this relationship may elucidate whether central arterial stiffness represents a clinically meaningful vascular biomarker that could inform risk stratification, comprehensive cognitive assessment, and future interventions targeting vascular brain health outcomes post-stroke.

Therefore, the purpose of this study was to examine the association between cfPWV and cognitive function, defined by Montreal Cognitive Assessment (MoCA) score ≤26/30, among individuals in the chronic phase of stroke recovery. The secondary objective was to evaluate the associations between cfPWV and MoCA cut points of ≤22/30 and ≤24/30 as additional cognitive impairment thresholds commonly used in stroke clinical practice.^32^ It is hypothesized that greater cfPWV would be associated with increased odds of cognitive impairment independent of demographic and cardiovascular risk factors.^31^

## Methods

This study was a retrospective cross-sectional analysis of baseline data collected from a multi-site RCT (NCT03614585).^33-35^ Research ethics approval was obtained (HIREB 4713, CRIR-1310-0218) and informed written consent was obtained from all participants. Enrollment occurred between January 2019 and August 2023. Methodological details for the RCT are reported elsewhere.^33-35^ The reporting of this study followed the STROBE guidelines for cross-sectional studies (Appendix S1). Deidentified participant data are available from the corresponding author upon reasonable request.

### Participants

Participants were recruited from the Jewish Rehabilitation Hospital and community members within the Montreal Metropolitan area, Quebec and the Hamilton, Ontario region. Enrollment occurred between January 2019 and August 2023. Participants were eligible for the larger trial if they were: between 40–80 years old and 6-60 months following the first-ever stroke confirmed by MRI/CT, living in the community, able to independently walk with or without a gait aid for at least 10 meters. Individuals were excluded from the trial if they: had a stroke of non-cardiogenic origin or tumor, scored > 2 on the modified Rankin Scale (e.g., defined as slight disability; unable to carry out all previous activities, but able to look after own affairs without assistance),^36^ had contraindications to cardiopulmonary exercise testing (CPET) or classified as class C or D American Heart Association Risk Criteria,^37^ were actively engaged in stroke rehabilitation services, had other neurological or musculoskeletal comorbidities, exhibited pain that worsened with exercise, or cognitive, communication, or behavioural issues that could limit their ability to provide consent or follow instructions. Participants with complete cfPWV and MoCA data at baseline were included the current analysis.

### Assessments

Demographic information, including age, biological sex, and medical history were collected for all participants at baseline. All assessments took place within one week. Information regarding timing post-stroke, stroke type, and stroke severity as assessed by the National Institutes of Health Stroke Severity Scale (NIHSS)^38^ were collected. Usual gait speed was evaluated over a 10-meter distance using their usual gait aid as appropriate. Walking capacity was evaluated using the 6-minute walk test (6MWT) following standardized guidelines using their usual gait aid as appropriate.^39^ The 6MWT was conducted in a 20-meter indoor hallway at baseline, and participants were instructed to walk as far as possible in 6 minutes, where the total distance walked in meters was recorded after the test. Cardiorespiratory fitness, defined as the peak oxygen uptake (mL/kg/min) was assessed using a symptom-limited CPET on recumbent stepper (NuStep T4r, NuStep LLC, Ann Arbor, MI, United States), following an adaptive incremental protocol validated for individuals with stroke.^40,41^

### Carotid-Femoral Pulse Wave Velocity (cfPWV)

All cfPWV assessments were conducted during one visit in temperature-controlled laboratories (∼23°C). Prior to the visit, participants were instructed to fast from food or drink for four hours, abstain from caffeine for eight hours and smoking for 12 hours, and to avoid structured physical activity for 24 hours. Participants were encouraged to adhere to routine medications as prescribed. Prior to cfPWV collection, participants were instrumented with a 3-lead electrocardiogram (Dual Bio Amp FE232, ADInstruments) to measure continuous heart rate over a 10-minute supine resting period. Blood pressure was collected from the brachial artery (Dinamap V100, General Electric Healthcare, Chicago, IL, United States Inc). Two blood pressure readings were measured and averaged and if values differed by>5 mmHg, 2 additional readings were taken and the average across all 4 readings was used.

Following the baseline resting period, cfPWV was assessed non-invasively using applanation tonometry (SPT-301, Millar Instruments Inc., Houston, TX, United States) connected to a pressure control unit (Millar Instruments Inc). Pressure signals were collected at 200 Hz using an analogue-to-digital data acquisition system (PowerLab, ADInstruments, Colorado Springs, CO, United States). Pressure waveforms were band-pass filtered (5-30 Hz) and the foot of each waveform was identified as the minimum value of the filtered signal (LabChart7 Pro, ADInstruments, Colorado Springs, CO, United States). Pulse waveforms were collected serially, one site at a time following the 10-minutes of supine rest. At least 30 consecutive and consistent pulse pressure waveforms were collected.^42^ cfPWV was calculated as the distance (meters)/Δt (seconds), where the distance was the measured using a straight and taut tape measure over the surface of the body between the carotid and femoral sites, and Δt represents the time delay between the foot of each waveform. cfPWV values were averaged across the 30 cardiac cycles and subsequently divided by 80% of the measured distance between the arterial sites.

### Global Cognitive Function

Global cognitive function was assessed using the (MoCA) and was the primary dependent variable of interest.^43^ The MoCA is a validated cognitive test for post-stroke cognitive impairment,^44^ with superior sensitivity to the Mini Mental State Exam (MMSE) for detecting vascular cognitive deficits in people with transient ischemic attack and stroke.^45^ The MoCA assesses multiple domains including attention, memory, visuospatial abilities, language, abstraction, concentration and orientation. The MoCA is an ordinal scale scored from 0 to 30, whereby lower scores are suggestive of lower cognitive performance, with a clinical categorical cut point score of <26/30 suggesting cognitive impairment.^43^ Recent evidence suggests using additional categorical cut point ranging from 22-25/30 may balance diagnostic accuracy for identifying post-stroke cognitive impairments.^32^

### Statistical Analyses

Participant demographic statistics were summarized as means ± standard deviation or medians (interquartile range [IQR]) for normally and non-normally distributed data, respectively, and frequencies (n, %) for categorical data. Baseline characteristics were disaggregated by sex to promote sex-based reporting in exercise stroke research.^46^ Differences in baseline characteristics between males and females were determined using Welch’s t-test with unequal variances, Wilcoxon rank-sum tests and Fisher’s exact tests for parametric, non-parametric and categorical data, respectively.

Logistic regression analyses were performed to examine the association between MoCA (primary dichotomous dependent variable at a MoCA cut point of ≤ 26/30) and cfPWV (primary independent continuous variable measured in m/s). The primary MoCA cut point of ≤26/30 was selected as this value has traditionally been associated with cognitive impairment.^43^ Secondary cut points of ≤24/30 and ≤22/30 were selected as additional sensitivity cut points since scores ranging between these scores are reported to balance diagnostic accuracy metrics for identifying post-stroke cognitive impairments.^32^

Unadjusted and adjusted logistic regression models were conducted and reported in the present manuscript. Models were first adjusted for demographic variables including covariates of age (years, continuous) and sex (male/female, binary). Models were then adjusted for clinical and cardiovascular risk factors including such as stroke severity (e.g., NIHSS), systolic blood pressure (mmHg, continuous), type-2 diabetes (present/absent, binary), and smoking history (ever smoker/non-smoker, binary). Covariate selection was driven by theory given their known associations on the primary and independent dependent variables.^13,14,47^ Requisite assumptions, including normality, residual and influencing outliers (e.g., Cook’s distance > 4/n), goodness of fit (e.g., Hosmer Lemeshow), and collinearity (e.g., variance inflation factor), were evaluated.

The area under the curve (AUC) and its precision estimates (95% CI) were used to evaluate the overall diagnostic accuracy of the logistic regression models, where AUC equals 0.5 when the Receiver Operating Curve (ROC) corresponds to random chance and 1.0 for perfect accuracy.^48^ AUC values are generally interpreted as no discrimination (AUC = 0.5), acceptable (AUC = 0.7 to 0.8), excellent (0.8 to 0.9), and perfect (0.9 to 1.0) discrimination.^49^ The level of significance was set to *p* = 0.05. All statistical analyses were conducted using R version 4.5.2.

## Results

Of 82 participants enrolled in the main trial, 68 participants with complete cfPWV and MoCA data at baseline were included in this analysis. Participant characteristics for the entire sample and disaggregated by sex are described in Table 1. Participants were on average 1.8 (standard deviation [SD] 1.2) years post-stroke and had mild to moderate stroke severity as assessed by the NIHSS (median = 1, IQR = 2). Males were older than females (mean difference [MD] = 5.2 years, p = 0.03). No other significant differences were observed between sexes, including cfPWV (p = 0.59), MoCA scores (p = 0.32), stroke type, stroke severity, cardiovascular risk factors, or physical function measures (Table 1). In total, 34 participants (50%) were identified as having a MoCA score ≤26, 21 (31%) with scores ≤24, and 15 (22%) with scores ≤22.

**Table 1.**
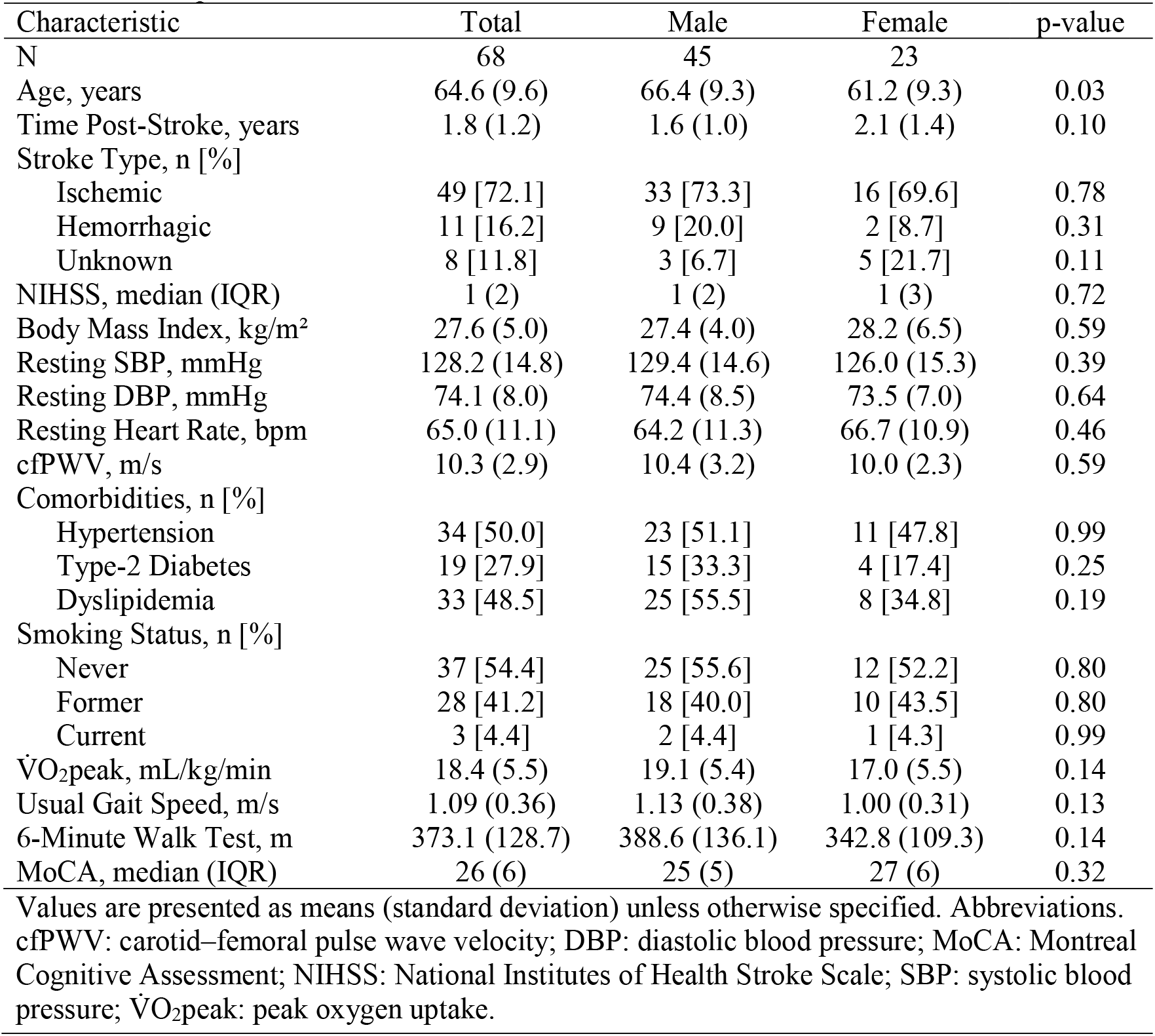
Participant characteristics.

Table 2 presents the unadjusted (Model 1), demographics-adjusted (Model 2: age, sex), and fully adjusted (Model 3: age, sex, systolic blood pressure, type-2 diabetes, NIHSS, and ever-smoker status) logistic regression analyses for each MoCA cut point. For the primary dependent variable, the unadjusted model for MoCA ≤26 demonstrated a significant association between cfPWV and cognitive impairment (Model 1; OR = 1.31; 95% CI 1.08, 1.64; p = 0.01; AUC = 0.68; 95% CI 0.55, 0.81). After adjusting for age and sex (Model 2), the association remained significant (OR = 1.32; 95% CI 1.06, 1.72; p = 0.02; AUC = 0.69; 95% CI 0.57, 0.82). In the fully adjusted model (Model 3, Figure 1 Panel A), each 1 m/s increase in cfPWV was associated with a 35% increase in the odds of having a MoCA score ≤26 (OR = 1.35; 95% CI 1.06, 1.81; p = 0.027; AUC = 0.77; 95% CI 0.66, 0.88; R^2^= 0.30, p-value = 0.014). There was no evidence of collinearity in the multivariable models (Model 2: VIF = 1.40; Model 3: VIF = 1.48) and the Hosmer-Lemeshow test indicated adequate model fit across all models (Model 1: χ^2^ = 6.51, p = 0.59; Model 2: χ^2^ = 9.18, p = 0.33; Model 3: χ^2^ = 6.05, p = 0.64). One influential observation was identified by Cook’s distance in the unadjusted model, two in Model 2, and five in Model 3, however these observations were deemed clinically plausible, did not influence the directionality of results, and were retained in the models to preserve the event counts per model. Sensitivity analyses with the removal of these influential observations are presented in Table 1 Appendix S2.

**Table 2.** Unadjusted and adjusted logistic regression models for each MoCA cutpoint.

| Variable | Model 1: Unadjusted |  |  | Model 2: Demographics |  |  | Model 3: Risk Factors |  |  |
| --- | --- | --- | --- | --- | --- | --- | --- | --- | --- |
|  | OR | 95% CI | p-value | OR | 95% CI | p-value | OR | 95% CI | p-value |
| 1) MoCA ≤ 26 (primary dependent cut point) | | | | | | | | | |
| cfPWV (m/s) | 1.31 | 1.08, 1.64 | 0.01 | 1.32 | 1.06, 1.72 | 0.02 | 1.35 | 1.06, 1.81 | 0.027 |
| Age (years) |  |  |  | 1.00 | 0.93, 1.06 | 0.94 | 0.99 | 0.92, 1.06 | 0.73 |
| Sex (female) |  |  |  | 0.51 | 0.16, 1.54 | 0.24 | 0.51 | 0.14, 1.74 | 0.29 |
| SBP (mmHg) |  |  |  |  |  |  | 1.01 | 0.97, 1.05 | 0.65 |
| T2D (present) |  |  |  |  |  |  | 1.43 | 0.41, 5.06 | 0.56 |
| NIHSS |  |  |  |  |  |  | 1.19 | 0.89, 1.69 | 0.26 |
| Ever Smoker |  |  |  |  |  |  | 3.73 | 1.23, 12.33 | 0.024 |
| 2) MoCA ≤ 24 (secondary dependent cut point) | | | | | | | | | |
| cfPWV (m/s) | 1.51 | 1.21, 1.96 | <0.001 | 1.36 | 1.05, 1.80 | 0.02 | 1.41 | 1.04, 2.01 | 0.037 |
| Age (years) |  |  |  | 1.10 | 1.02, 1.22 | 0.03 | 1.11 | 1.01, 1.26 | 0.045 |
| Sex (female) |  |  |  | 1.73 | 0.46, 6.78 | 0.42 | 2.40 | 0.51, 12.22 | 0.27 |
| SBP (mmHg) |  |  |  |  |  |  | 1.03 | 0.98, 1.09 | 0.20 |
| T2D (present) |  |  |  |  |  |  | 2.27 | 0.45, 12.56 | 0.32 |
| NIHSS |  |  |  |  |  |  | 1.44 | 1.05, 2.18 | 0.039 |
| Ever Smoker |  |  |  |  |  |  | 2.87 | 0.70, 13.62 | 0.15 |
| 3) MoCA ≤ 22 (secondary dependent cut point) | | | | | | | | | |
| cfPWV (m/s) | 1.37 | 1.11, 1.77 | 0.006 | 1.31 | 1.04, 1.72 | 0.03 | 1.33 | 1.03, 1.79 | 0.039 |
| Age (years) |  |  |  | 1.04 | 0.96, 1.13 | 0.39 | 1.03 | 0.94, 1.14 | 0.54 |
| Sex (female) |  |  |  | 0.86 | 0.20, 3.40 | 0.84 | 0.77 | 0.13, 3.75 | 0.75 |
| SBP (mmHg) |  |  |  |  |  |  | 1.03 | 0.98, 1.09 | 0.26 |
| T2D (present) |  |  |  |  |  |  | 1.42 | 0.28, 6.80 | 0.65 |
| NIHSS |  |  |  |  |  |  | 1.38 | 1.01, 1.96 | 0.048 |
| Ever Smoker |  |  |  |  |  |  | 2.61 | 0.64, 12.23 | 0.19 |

Abbreviations: MoCA: Montreal Cognitive Assessment; NIHSS: National Institute of Health Stroke Severity; OR: Odds Ratio; PWV: pulse wave velocity; SBP: systolic blood pressure; T2D: Type-2 diabetes.

**Figure 1.**
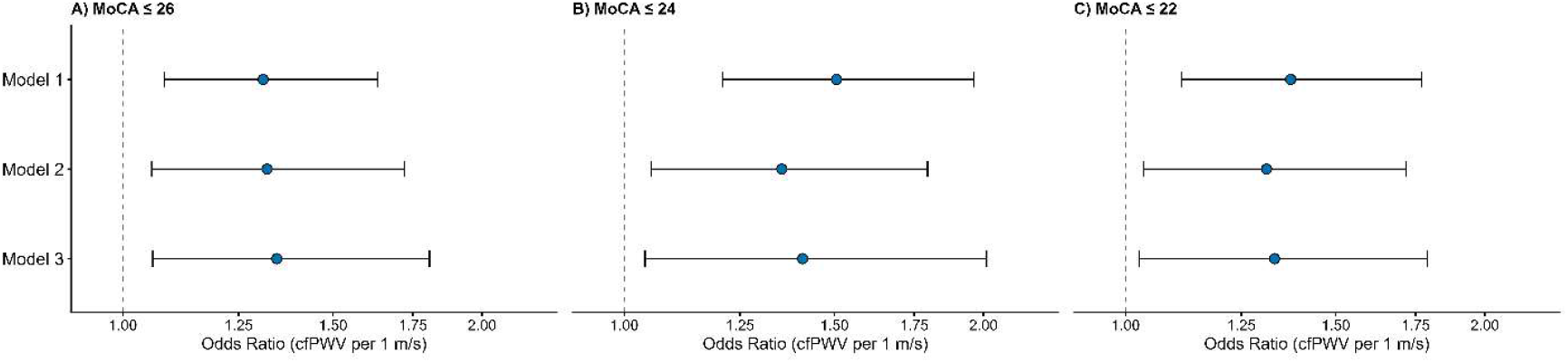
Adjusted odds ratios (95% CI) for cognitive impairment per 1 m/s increase in cfPWV at MoCA cutpoints of ≤26 (A), ≤24 (B), and ≤22 (C). Model 1, unadjusted; Model 2, adjusted for age and sex; Model 3, adjusted for age, sex, systolic blood pressure, type 2 diabetes, NIHSS, and ever-smoker status. Dashed line indicates OR = 1.0.X-axis is presented on a log scale.

For the secondary cut points, unadjusted and demographics-adjusted models for MoCA ≤24 and MoCA ≤22 demonstrated consistent significant associations between cfPWV and cognitive impairment (Table 2). In the fully adjusted model (MoCA ≤24, Model 3, Figure 1 Panel B), each 1 m/s increase in cfPWV was associated with a 41% increase in the odds of having a MoCA score ≤24 (OR = 1.41; 95% CI 1.04, 2.01; p = 0.037; AUC = 0.88; 95% CI 0.79, 0.98; R^2^= 0.51, p-value < 0.001). Similarly, in the fully adjusted model (MoCA ≤22, Model 3, Figure 1 Panel C), each 1 m/s increase in cfPWV was associated with a 33% increase in the odds of having a MoCA score ≤22 (OR = 1.33; 95% CI 1.03, 1.79; p = 0.039; AUC = 0.82; 95% CI 0.68, 0.96, R^2^= 0.35, p-value =0.013). Four to eight influential observations were identified by Cook’s distance in these models; however, these observations were deemed clinically plausible, did not influence the directionality of results, and were retained in the models to preserve the event counts per model. The results of sensitivity analyses accounting for these influential observations are presented in Table 1 Appendix S2.

## Discussion

This study demonstrated that gold-standard cfPWV was positively and significantly associated with post-stroke cognitive impairment, as measured by the MoCA, among individuals in the chronic phase of stroke recovery. Importantly, this association was consistent across all three MoCA cutpoints (≤26, ≤24, and ≤22) and remained consistent after sequential adjustment for demographic variables (age, sex), stroke severity, and established cardiovascular risk factors (systolic blood pressure, type-2 diabetes, and ever-smoker status). Across adjusted models, each 1 m/s increase in cfPWV was associated with 1.33 to 1.41 higher odds of post-stroke cognitive impairment. Further, the fully adjusted models demonstrated acceptable to excellent discrimination (AUC 0.77 to 0.88), with the strongest model performance observed at the MoCA ≤24 cutpoint. These findings are notable given that each 1 m/s increase in aortic PWV has been associated with a 14% higher adjusted risk of total cardiovascular events and a 15% higher adjusted risk of cardiovascular or all-cause mortality.^50^ In the context of cognitive impairment, each 1 m/s increase in estimated PWV was associated with a 51% higher adjusted rate of incident dementia and a 58% higher adjusted rate of Alzheimer’s disease.^51^ Taken together, our findings suggest that cfPWV, the gold-standard measure of arterial stiffness,^18^ may be a clinically relevant vascular biomarker associated with commonly used MoCA thresholds for post-stroke cognitive impairment. cfPWV may therefore provide complementary information about vascular aging and cerebrovascular risk after stroke that may not be captured by traditional cardiovascular risk assessment or standardized cognitive screening.

Our findings are consistent with previously reported associations between aortic stiffness and cognitive outcomes in non-stroke populations. Higher cfPWV has been prospectively associated with a 41% increased risk of incident mild cognitive impairment over a median 10-year follow-up, following adjustment for age, sex and cardiovascular risk factors.^28^ This relationship was further supported by a recent systematic review and meta-analysis of 52 studies in non-stroke populations which found that elevated cfPWV was associated with poor cognitive performance across global cognition, executive function, and memory.^17^ To our knowledge, only one prior study has examined this relationship among individuals post-stroke, which found an inverse association between cfPWV and MMSE, but the association was attenuated after adjustment for confounding variables, including handgrip strength, static balance, walking distance and cardiovascular risk factors.^31^ The lack of association in this previous work may also partly reflect the limited cognitive impairment of the sample, in which most participants scored in the cognitively normal range (MMSE ≥ 24; mean 26 ± 2), as well as the limited sensitivity of the MMSE for detecting mild cognitive impairment post-stroke.^45^ Our findings extend this work by including a broader and more representative sample of individuals post-stroke, where approximately 50% of the sample were identified as cognitively impaired using the MoCA ≤ 26 cut point, and by using the MoCA which has superior sensitivity to the MMSE for detecting vascular cognitive deficits after stroke.^45^ The consistency of our findings across clinically referenced MoCA cut points, in addition to sequential adjustment for confounding demographic and cardiovascular risk factors and inclusion of stroke severity in the statistical models, provides further support for an association between cfPWV and post-stroke cognitive impairment. Indeed, future longitudinal studies in stroke are warranted to evaluate whether novel biomarkers like cfPWV improves prognostic risk stratification when combined with standardized cognitive testing, traditional cardiovascular risk factors, and neuroimaging biomarkers.^11^

There are possible mechanisms that may explain the association between cfPWV and cognitive impairment. Aging is accompanied by vascular remodeling and progressive stiffening of central elastic arteries,^52^ which reduces their buffering capacity and allows for greater pulsatile energy that is transmitted to the cerebrovasculature.^53^ Increased arterial stiffness and pulsatile flow to the brain has been associated with reduced cerebral blood flow, white and gray matter damage,^54,55^ and blood-brain barrier disruption,^15^ all of which are factors that contribute to cognitive decline.^52,56^ In people post-stroke, these mechanisms are particularly relevant since the brain may already be vulnerable due to the index stroke and co-existing cerebrovascular disease.^11^ Notably, cfPWV values in the present sample were high (10.4 m/s in males and 10.0 m/s in females) and corresponded to approximately the 90th percentile relative to age- and sex-matched reference values.^57^ Thus, elevated cfPWV may reflect not only vascular aging, but also ongoing hemodynamic stress on a pre-existing compromised vascular bed, which could plausibly contribute to post-stroke cognitive impairment, and in some cases, progression towards post-stroke dementia.^58^ While stroke-specific vascular mechanisms and cognitive impairment remain an emerging area of research,^11^ these findings suggest that elevated cfPWV may represent a clinically relevant biomarker of cognitive impairment after stroke that may be amenable to change following targeted interventions.

There is currently limited pharmacological treatment for post-stroke cognitive impairment and dementia.^11^ Evidence suggests that cholinesterase inhibitors may improve outcomes of cognition, however, this evidence is specific to patients with various types of vascular dementia.^59^ From a non-pharmacological perspective, exercise training appears to be a promising intervention to improve both arterial stiffness^60^ and cognitive function in non-stroke populations.^61^ In people post-stroke, high-intensity exercise appears to provide the greatest cardiovascular benefit,^62^ including improvements in cardiorespiratory fitness and systolic blood pressure, both of which may be determinants of cfPWV^63^ and cognitive function.^64^ However, a recent trial reported no significant effect of either high-intensity interval training or moderate intensity continuous training on cfPWV or systolic blood pressure^34^, although it was not statistically powered to detect changes in either outcome. Evidence also suggests that exercise may improve cerebrovascular outcomes^65^ that may underpin changes in cognitive function,^52,66^ such as changes in cerebral blood velocity^67^ and cerebral blood flow.^68,69^ Taken together, these findings suggest that exercise training could influence both vascular and cognitive health after stroke, but the extent to which these benefits are mechanistically linked remains unclear. Further research is needed to determine whether exercise-induced improvements in vascular function, including reductions in cfPWV or improvements in cerebral blood flow, are associated with meaningful improvements in cognitive performance post-stroke.

It is also important to highlight the limitations of the current study. The cross-sectional design does not allow for the inference of causal pathways between cfPWV and post-stroke cognitive impairment. Second, while the current analysis accounted for stroke severity, the study sample only included participants with mild to moderate severity, which may limit the generalizability to those with more severe disability who may be at highest risk for post-stroke cognitive impairment. Lastly, we used the MoCA, which is a global cognitive outcome that although is used clinical clinically and more sensitive to the MMSE,^45,70^ it may not be as sensitive to capture changes in cognitive assessment compared to more comprehensive cognitive batteries.^71^ As such, the model estimates and the AUC values should be interpreted as model discrimination within this sample of stroke survivors. Indeed, future work is required to confirm our findings using larger studies that leverage longitudinal designs to evaluate possible causal pathways between cfPWV and cognitive function as assessed using comprehensive cognitive batteries.

In conclusion, higher cfPWV was consistently associated with greater odds of post-stroke cognitive impairment across clinically referenced MoCA cut points after adjustment for demographic factors, stroke severity, and cardiovascular risk factors. These findings support that cfPWV may be a complementary vascular biomarker of cognitive impairment in people with mild to moderate stroke severity. Future longitudinal studies are needed to determine whether cfPWV improves prognostic risk stratification and helps identify individuals who may benefit from closer cognitive monitoring, cardiovascular risk-factor management, or vascular-targeted interventions after stroke.

## Data Availability

Deidentified participant data are available from the corresponding author upon reasonable request.

## Acknowledgements

The authors would like to thank Hanna Fang for the coordination of this study.

## Sources of Funding

This study has been funded by an operating grant from the Canadian Institute of Health Research (388320). Kevin Moncion and Lynden Rodrigues are both supported by a Canadian Institute of Health Research (CIHR) Postdoctoral Fellowship. Juliano Abreu and Adam Sutoski are supported by a CIHR Graduate Scholarship. Marc Roig is supported by a Salary Award (Junior II) from Fonds de Recherche Santé Québec (FRQS).

## Notes

### Competing Interest Statement

The authors have declared no competing interest.

### Clinical Trial

This study was a retrospective cross-sectional analysis of baseline data collected from a multi-site RCT (NCT03614585)

### Author Declarations

Research ethics approval was obtained (HIREB 4713, CRIR-1310-0218) and informed written consent was obtained from all participants. Hamilton Integrated Research Ethics Board (HIREB). HiREB represents the institutions of Hamilton Health Sciences, St. Joseph's Healthcare Hamilton, Research St. Joseph's-Hamilton, and the Faculty of Health Sciences at McMaster University in Hamilton Ontario, Canada. Centre de recherche interdisciplinaire en readaption du Montreal metropolitain (CRIR), CISSS de Laval, Jewish Rehabilitation Hospital (JRH), Laval, Quebec, Canada.

